# Longitudinal Analysis of CYFRA 21-1 Levels in Patients with Pulmonary Nodules: Differential Trajectories Between Benign and Malignant Cases and Impact of Tumor Resection

**DOI:** 10.64898/2026.01.10.26343848

**Authors:** Yency J. Forero, Michael N. Kammer, Kevin C. McGann, Sheau-Chiann Chen, Heidi Chen, Samson Argaw, Timothy A. Khalil, Sanja L. Antic, Yong Zou, Zuo Lianrui, Thomas A. Lasko, Bennet A. Landman, Stephen A. Deppen, Eric L. Grogan, Fabien Maldonado

## Abstract

**Background:** CYFRA 21-1, a cytokeratin-19 fragment, is a validated serum biomarker for non-small cell lung cancer (NSCLC). However, most studies rely on single time-point measurements, limiting its specificity in differentiating malignancy from benign pulmonary conditions. Inspired by the clinical utility of serial PSA measurements in prostate cancer, we investigated whether longitudinal trends in CYFRA 21-1 could enhance diagnostic and monitoring capabilities in patients with pulmonary nodules

**Methods and Findings:** We analyzed 132 patients with pulmonary nodules, including 41 with lung cancer and 91 with benign diagnoses. CYFRA 21-1 levels were measured serially using electrochemiluminescence assays. Longitudinal trends were assessed using linear mixed-effects models to estimate biomarker trajectories. Subgroup analyses examined differences between benign, untreated cancer, and post-treatment cancer groups, as well as within-patient changes in a subset of 16 cancer patients with both pre- and post-surgical measurements. Log-transformed data were used for the analysis. At baseline, CYFRA 21-1 levels were significantly higher in malignant versus benign nodules. Over time, CYFRA trajectories diverged: benign cases showed slight increases, whereas cancer patients exhibited greater biomarker volatility. In treated cancer patients, trend of CYFRA levels on the natural log scale decline from –0.00137 pre-surgery to – 0.00263 to post-surgery, and both cancer groups showed significantly higher absolute slopes than the benign group (p < 0.05). While pre- vs post-treatment slope differences did not reach significance (p = 0.211), the general pattern indicated that CYFRA 21-1 is a dynamic marker responsive to tumor presence and removal.

**Conclusions:** CYFRA 21-1 exhibits substantial within-patient variability over time, with trajectories that reflect disease state and treatment. These findings suggest that longitudinal monitoring of CYFRA 21-1—analogous to PSA velocity in prostate cancer— may offer improved diagnostic and prognostic insight in the evaluation of pulmonary nodules. Further studies in larger cohorts are warranted to validate these findings and explore clinical implementation of CYFRA trajectory analysis.

## Introduction

Cytokeratin-19 fragment (CYFRA 21-1) is a well-established serum biomarker that has been widely studied in lung cancer, particularly non-small cell lung cancer (NSCLC), for both diagnostic and prognostic purposes^1, 2^. Many studies have evaluated CYFRA 21-1 for diagnosis or prognosis of lung cancer, either alone or in combination with other biomarkers^3–7^. However, most investigations of CYFRA 21-1 focus on single time-point measurements (e.g. baseline levels) rather than how the biomarker changes over time within individual patients. This leads to poor specificity, given CYFRA 21-1’s elevated levels because of inflammation or other epithelial comorbidities, such as Chronic Obstructive Pulmonary Disease (COPD) and Interstitial Lung Fibrosis (IPF^)8^.

In contrast, the clinical management of prostate cancer has long recognized the importance of biomarker trajectories. Prostate-specific antigen (PSA) “velocity” – the rate of PSA change – is an established indicator for malignancy and disease progression. For example, a rise in PSA greater than ∼0.75 ng/mL per year is considered suspicious for prostate cancer, even if absolute PSA values are not yet above a static threshold^9^. Serial PSA measurements (velocity and doubling time) are routinely used to trigger biopsies, guide treatment decisions, and monitor for recurrence. This analogy suggests that tracking the temporal trajectory of a cancer biomarker can provide critical information beyond a single snapshot.

By extension, the trajectory of CYFRA 21-1 might enhance the evaluation of pulmonary nodules. Pulmonary nodules are a common clinical dilemma, requiring differentiation between benign lesions and early lung cancers. While a single elevated CYFRA 21-1 level can support a lung cancer diagnosis, longitudinal changes in CYFRA 21-1 could potentially signal malignancy (or benign behavior) earlier or more reliably. Recent research in lung cancer screening has begun exploring serial biomarker algorithms: repeated measurements of panels including CYFRA 21-1 have shown improved sensitivity and earlier detection of lung cancer compared to a one-time threshold approach^10^. Despite this interest, the temporal behavior of CYFRA 21-1 within individual patients – for example, whether CYFRA trends upward in growing cancers or remains stable in benign nodules – remains understudied.

Here we present a pilot longitudinal analysis of CYFRA 21-1 in patients with pulmonary nodules, comparing the trajectories in those with benign vs. malignant outcomes. We also examine how surgical removal of a malignant nodule (tumor resection) affects the CYFRA trajectory within the same patient. Our approach parallels the PSA velocity concept, positing that dynamic changes in CYFRA 21-1 over time may offer diagnostic and monitoring value. We hypothesize that malignant nodules will exhibit distinct CYFRA 21-1 kinetics (such as rising levels over time) compared to benign nodules, and that effective treatment (surgery) will alter the biomarker’s course (for example, causing a decline). This manuscript describes our cohort and methods and presents initial findings that underscore the potential importance of CYFRA 21-1 trajectories in pulmonary nodule assessment.

## Methods

### Study Cohort

This prospective collection with retrospective blinded evaluation (PROBE) study analyzed serum samples from a cohort of patients with pulmonary nodules who underwent serial blood draws over the course of their clinical evaluation. Serum was processed and stored within 2 hours of blood draw, according to the Early Detection Research Network’s Lung Clinical Validation Center standard operating protocol^11^. All patients were eventually categorized as having either benign nodules or malignant nodules (lung cancer) based on definitive diagnoses (e.g. histopathology or long-term imaging follow-up).

### CYFRA 21-1 Measurement

Samples were obtained from the VUMC Thoracic Biorepository11 and accessed for research purposes between 01/01/2008 to 31/12/2018. Serum CYFRA 21-1 concentrations were measured using the Roche Elecsys Cobas e411 analyzer (electrochemiluminescence immunoassay), following the manufacturer’s protocol, with a College of American Pathologist (CAPP) certified laboratory. All samples were run concurrently, in a randomized order and in a blind fashion.

### Statistical Analysis

We utilized linear mixed-effects models (LME) to analyze longitudinal trends in CYFRA 21-1 while accounting for repeated measures within patients. Main effect of time, cancer group and their interaction as the fixed effect and patient ID as a random intercept (to account for baseline differences between individuals). This allowed estimation of group-specific trends while accounting for repeated measures within individuals. CYFRA values were log-transformed for modeling, due to the right-skewed distribution of raw concentrations. “Group” in the model was defined with three categories: *Benign*, *Cancer– Untreated*, and *Cancer–Treated*. Here, “Cancer–Untreated” refers to measurements taken while a known cancer nodule was still present (pre-surgery), and “Cancer–Treated” refers to measurements taken after surgical removal of the cancer. Time was measured as the duration from each patient’s first CYFRA measurement (baseline) to the current measurement (in days). We fit the LME model by maximum likelihood and performed an F-test (type III ANOVA) to assess the significance of fixed effects (in particular, to test whether there were overall differences between groups and whether time trends differed between groups via the interaction). Post-hoc pairwise comparisons of estimated marginal means were conducted via Wald test using the emmeans package, with Tukey adjustment, to compare mean CYFRA levels between groups on the log scale and then reported on the original scale with 95% confidence intervals. To assess differences in CYFRA 21-1 trajectories across diagnostic groups, s absolute slope of log-transformed CYFRA values by groups was performed using one-way ANOVA analysis, where the slope was first estimated using the **lmList** function from the **nlme** R package, and then the absolute value of each slope was calculated. The P-value was adjusted using Tukey method for comparing a family of 3 estimates.

In addition, we specifically evaluated paired changes in CYFRA 21-1 for the subset of cancer patients who had both a pre-surgery and post-surgery measurement. For each of these patients, the last CYFRA value before surgery was compared to the first CYFRA value after surgery. Due to the non-normal distribution of data, Wilcoxon signed-rank test was used to assess if there was a systematic change in CYFRA levels following tumor resection. This test assesses whether the median of the paired differences (post minus pre) is zero or not. All statistical analyses were performed in R (v4.4.0), with α = 0.05 as the significance threshold (two-tailed).

### Ethical considerations

The study was conducted according to the Declaration of Helsinki in its current version and was approved by the Institution Board Review (IRB) of Vanderbilt University Medical Center (IRB#030763). Written informed consent was obtained from all participants, and all methods were carried out in accordance with local law and regulations.

## Results

Among the 132 patients analyzed, 41 had malignant pulmonary nodules and 91 had benign nodules. Groups were similar in age, sex, race, smoking status, pack-years and lung nodule location (p > 0.05). However, history of previous cancer (21 of 41 malignant vs. 28 of 91 benign, p = 0.024), lung cancer history (5 of 41 malignant vs. 2 of 91 benign, p = 0.03), and histology diagnosis (p < 0.001) differed significantly between groups. The mean age was similar between groups (benign 63.2 ± 4.7 years vs malignant 64.6 ± 7.0 years). In the benign group, 57 were male and 34 were female; in the malignant group, 25 were male and 16 were female. Nearly all patients were Caucasian (85 of 91 in the benign group; 41 of 41 in the malignant group). Ever-smoking was nearly universal (91 of 91 benign; 39 of 41 malignant), with mean pack-years of 53.7 ± 23.7 for benign and 57.7 ± 32.2 for malignant patients.

Lung nodules varied: in were the right upper lobe (RUL) 20 benign and 10 malignant; in the right lower lobe (RLL), were 13 benign and 10 malignant. Malignant cases had higher proportions of central lesions (e.g., right hilum, 3 cases). Histology for malignant nodules included 23 adenocarcinomas, 11 squamous cell carcinomas, and samples numbers of other NSCLC subtypes or non-lung primaries. Benign nodules included 73 normal tissue samples, 12 negatives for malignancy, and a small number with metaplasia or unsatisfactory samples. **(Table 1)**

**Table 1.**
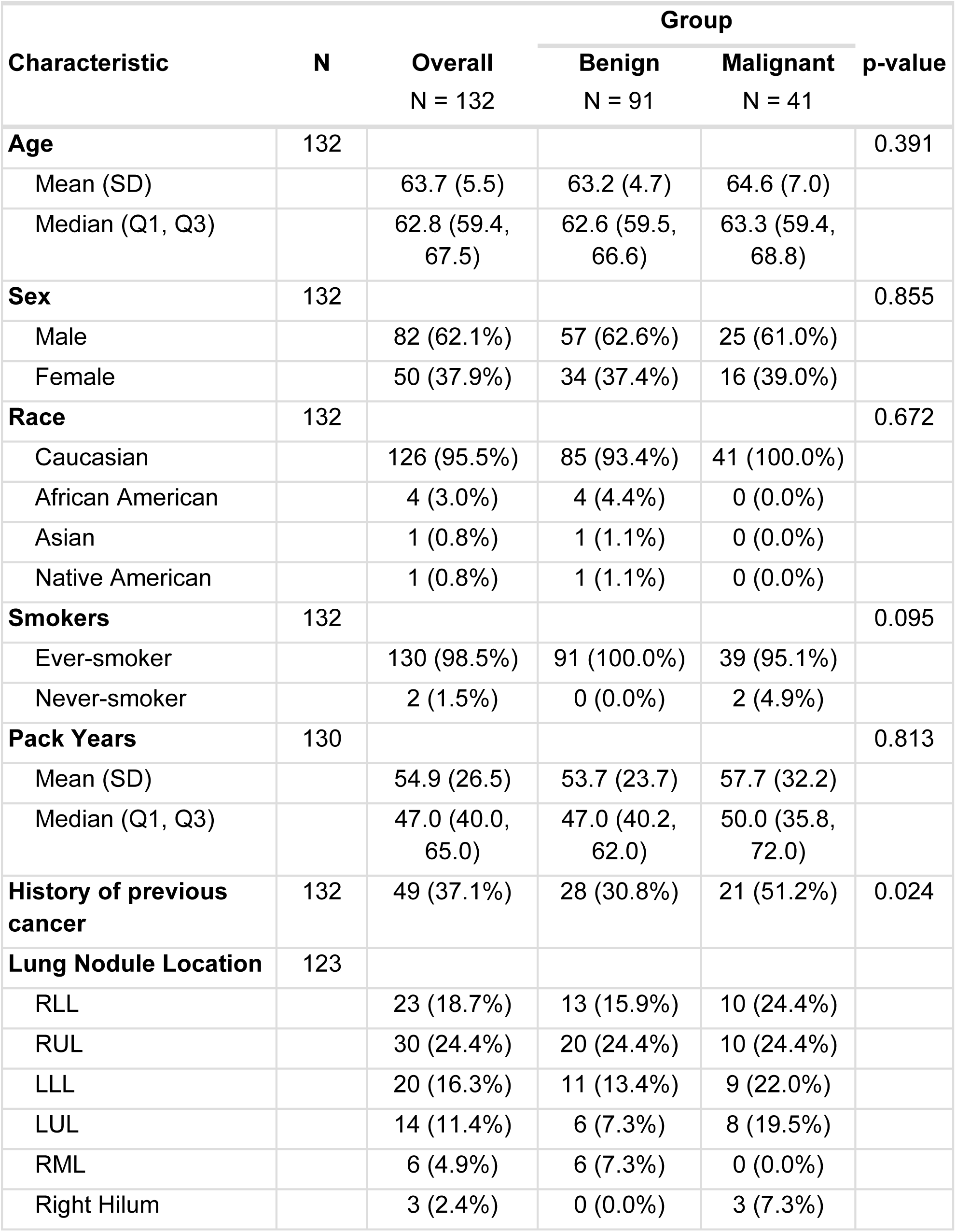
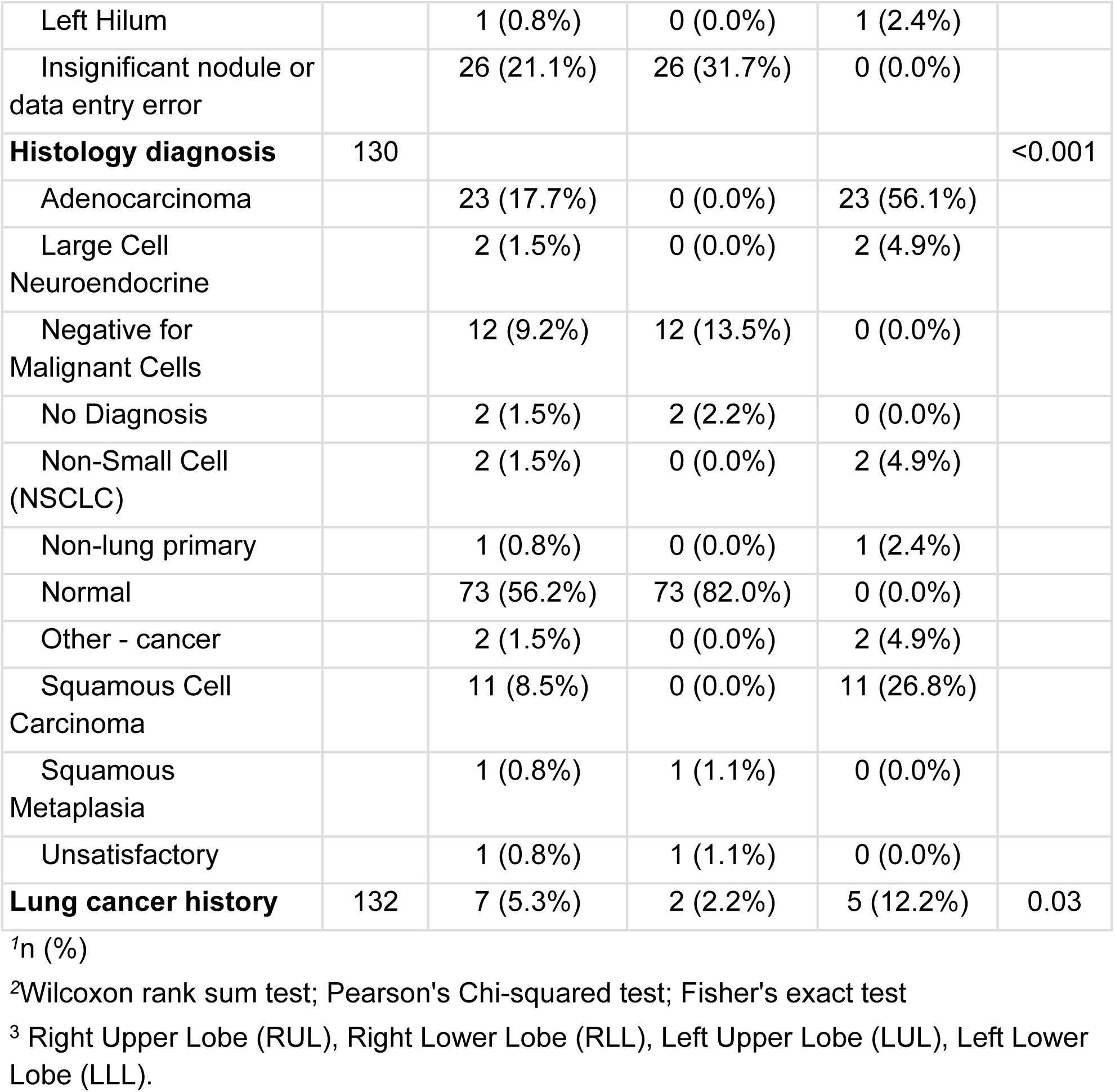
Summary of Patients Characteristics.

| Characteristic | N | Overall<br>N = 132 | Group |  | p-value |
| --- | --- | --- | --- | --- | --- |
|  |  |  | Benign<br>N = 91 | Malignant<br>N = 41 |  |
| **Age** | 132 |  |  |  | 0.391 |
| Mean (SD) |  | 63.7 (5.5) | 63.2 (4.7) | 64.6 (7.0) |  |
| Median (Q1, Q3) |  | 62.8 (59.4, 67.5) | 62.6 (59.5, 66.6) | 63.3 (59.4, 68.8) |  |
| **Sex** | 132 |  |  |  | 0.855 |
| Male |  | 82 (62.1%) | 57 (62.6%) | 25 (61.0%) |  |
| Female |  | 50 (37.9%) | 34 (37.4%) | 16 (39.0%) |  |
| **Race** | 132 |  |  |  | 0.672 |
| Caucasian |  | 126 (95.5%) | 85 (93.4%) | 41 (100.0%) |  |
| African American |  | 4 (3.0%) | 4 (4.4%) | 0 (0.0%) |  |
| Asian |  | 1 (0.8%) | 1 (1.1%) | 0 (0.0%) |  |
| Native American |  | 1 (0.8%) | 1 (1.1%) | 0 (0.0%) |  |
| **Smokers** | 132 |  |  |  | 0.095 |
| Ever-smoker |  | 130 (98.5%) | 91 (100.0%) | 39 (95.1%) |  |
| Never-smoker |  | 2 (1.5%) | 0 (0.0%) | 2 (4.9%) |  |
| **Pack Years** | 130 |  |  |  | 0.813 |
| Mean (SD) |  | 54.9 (26.5) | 53.7 (23.7) | 57.7 (32.2) |  |
| Median (Q1, Q3) |  | 47.0 (40.0, 65.0) | 47.0 (40.2, 62.0) | 50.0 (35.8, 72.0) |  |
| **History of previous cancer** | 132 | 49 (37.1%) | 28 (30.8%) | 21 (51.2%) | 0.024 |
| **Lung Nodule Location** | 123 |  |  |  |  |
| RLL |  | 23 (18.7%) | 13 (15.9%) | 10 (24.4%) |  |
| RUL |  | 30 (24.4%) | 20 (24.4%) | 10 (24.4%) |  |
| LLL |  | 20 (16.3%) | 11 (13.4%) | 9 (22.0%) |  |
| LUL |  | 14 (11.4%) | 6 (7.3%) | 8 (19.5%) |  |
| RML |  | 6 (4.9%) | 6 (7.3%) | 0 (0.0%) |  |
| Right Hilum |  | 3 (2.4%) | 0 (0.0%) | 3 (7.3%) |  |
| Left Hilum |  | 1 (0.8%) | 0 (0.0%) | 1 (2.4%) |  |
| Insignificant nodule or data entry error |  | 26 (21.1%) | 26 (31.7%) | 0 (0.0%) |  |
| **Histology diagnosis** | 130 |  |  |  | <0.001 |
| Adenocarcinoma |  | 23 (17.7%) | 0 (0.0%) | 23 (56.1%) |  |
| Large Cell Neuroendocrine |  | 2 (1.5%) | 0 (0.0%) | 2 (4.9%) |  |
| Negative for Malignant Cells |  | 12 (9.2%) | 12 (13.5%) | 0 (0.0%) |  |
| No Diagnosis |  | 2 (1.5%) | 2 (2.2%) | 0 (0.0%) |  |
| Non-Small Cell (NSCLC) |  | 2 (1.5%) | 0 (0.0%) | 2 (4.9%) |  |
| Non-lung primary |  | 1 (0.8%) | 0 (0.0%) | 1 (2.4%) |  |
| Normal |  | 73 (56.2%) | 73 (82.0%) | 0 (0.0%) |  |
| Other - cancer |  | 2 (1.5%) | 0 (0.0%) | 2 (4.9%) |  |
| Squamous Cell Carcinoma |  | 11 (8.5%) | 0 (0.0%) | 11 (26.8%) |  |
| Squamous Metaplasia |  | 1 (0.8%) | 1 (1.1%) | 0 (0.0%) |  |
| Unsatisfactory |  | 1 (0.8%) | 1 (1.1%) | 0 (0.0%) |  |
| **Lung cancer history** | 132 | 7 (5.3%) | 2 (2.2%) | 5 (12.2%) | 0.03 |

<sup>1</sup>n (%)
<sup>2</sup>Wilcoxon rank sum test; Pearson's Chi-squared test; Fisher's exact test
<sup>3</sup> Right Upper Lobe (RUL), Right Lower Lobe (RLL), Left Upper Lobe (LUL), Left Lower Lobe (LLL).

Among the 41 malignant cases, 16 patients underwent surgical resection and were analyzed as the surgical subgroup. The mean age of these patients was similar to the overall malignant group. Histological diagnoses were predominantly non-small cell lung cancer subtypes, including 10 adenocarcinomas and 5 squamous cell carcinomas, with 1 large cell neuroendocrine carcinoma. Clinical staging at diagnosis ranged from stage IA to IIIB, reflecting the diversity in disease extent among surgical candidates. Pathologic staging similarly showed heterogeneity with many early-stage resections but also some locally advanced cases.

Time to event analysis for the surgical subgroup showed a wide range of outcomes. Several patients had prolonged survival exceeding 60 months from diagnosis, while others experience recurrence or death within 24 months. The median time to event was approximately 39 months. These data highlight the clinical heterogeneity of resected lung cancers, supporting the need for individualized prognosis and biomarker assessment. **(Table 2)**

**Table 2.** Clinical, pathological and outcome characteristics of patients with confirmed malignancy and who underwent surgical resection.

| Clinical Stage | Pathologic Stage | Histology | Location | Metastasis | Cancer History | Survival Status | Time to Event (months) (death, alive or last follow up) |
| --- | --- | --- | --- | --- | --- | --- | --- |
| IIIA | T2N2M0 | Squamous cell carcinoma | LLL | No | No | Unknown | 12 |
| IIIA | T2N2M0 | Adenocarcinoma | LLL | Yes (CNS) | No | Deceased | 15 |
| IB | T1N0M0 | Squamous cell carcinoma | LUL | Yes (bone) | No | Deceased | 77 |
| IIA | T1aN1M0 | Adenocarcinoma | RLL | Yes (CNS) | No | Alive | 104 |
| IIA | T1aN1M0 | Adenocarcinoma | LUL | Yes (hilar) | No | Deceased | 24 |
| IB | T2aN0M0 | Squamous cell carcinoma | RLL | Yes (hilar) | No | Deceased | 91 |
| IA | T1bN0M0 | Neuroendocrine | LLL | No | No | Alive | 77 |
| IIIA | T3N1M0 | Adenocarcinoma | RLL | Unknown | No | Deceased | 39 |
| IB | T1bN0M0 | Adenocarcinoma | LUL | Unknown | No | Unknown | 80 |
| IIIA |  | Squamous cell carcinoma | Lymph node(4R) | Yes (bone) | Yes (colon) | Deceased | 38 |
| IIB | T3N0M0 | Adenocarcinoma | RLL | No | No | Unknown | 37 |
| IIA | T1bN0M0 | Neuroendocrine | RUL | No | No | Alive | 101 |
| N/A | N/A | Metastatic adenocarcinoma | LUL | Yes | Yes (Ampullar adenocarcinoma) | Deceased | 26 |
| IIIB | T4N3M0 | Adenocarcinoma | LLL | Yes | No | Unknown | 29 |
| IIB | T3N0M0 | Squamous cell carcinoma | RUL | No | No | Unknown | 4 |

Note: Follow-up information was unavailable for patient 16.
None of the surgically resected patients received Stereotactic Body Radiation Therapy (SBRT) for the resected pulmonary nodule. Data on SBRT used for metastatic sites during follow-up were not systematically collected.

None of the benign cases underwent surgical removal of the nodule during the study; these patients were followed with serial observations alone. By contrast, the malignant cases in the surgical subgroup had detailed staging information and contributed to the serial CYFFRA 21-1 measurements both before and after surgical resection of the tumor, providing internal “pre vs post” comparisons. Five of the cancer patients had only a single CYFRA measurement (at the initial visit, with no follow-up sample – these were patients who, for example, were lost to follow-up or had immediate treatment elsewhere). The remaining cancer patients had multiple serial measurements in at least one phase (pre- or post-treatment). The median follow-up duration for biomarker measurements was approximately 6–12 months, with a median of 3 blood draws per patient (range 1 to 9, as some patients in the paired group had frequent follow-up draws). Notably, CYFRA 21-1 levels were generally low in benign patients across all time points, though one benign-case patient exhibited an outlier high value (peak CYFRA 11.3 ng/mL) despite ultimately having a benign diagnosis. This was an exceptional case; most benign nodules had CYFRA levels well below the typical diagnostic cut-off of 3.3 ng/mL.

### Baseline CYFRA Levels (Benign vs Malignant)

At the initial measurement (baseline), serum CYFRA 21-1 levels were significantly higher in patients with malignant nodules compared to those with benign nodules.

### Longitudinal CYFRA levels (Benign vs Malignant)

The LME model’s estimated overall marginal means (back-transformed from log scale) indicate that, before any treatment, lung cancer patients had an average CYFRA level of 2.80 ng/mL (95% confidence interval 2.30–3.41), whereas benign nodule patients had an average of 1.87 ng/mL (95% CI 1.71–2.04). This difference (representing as a ratio of benign patients to cancer patients of 0.668 because tests were performed on the log scale) was statistically significant (p < 0.001). The group difference remained significant even for cancer patients *after* surgical removal of the nodule (a ratio of benign patients to cancer patients with post-resection of 0.711): the mean CYFRA in the post-resection state was 2.63 ng/mL (95% CI 2.21–3.12), which was still higher than the benign group (p = 0.002). There was no significant difference between the untreated-cancer and treated-cancer groups’ means (2.80 vs 2.63, respectively; p = 0.74), suggesting that, on average, CYFRA levels in patients shortly after surgery were slightly lower than pre-surgery levels but with considerable overlap. These comparisons reinforce that CYFRA 21-1 is typically elevated in lung cancer patients compared to those with benign nodules, consistent with its known diagnostic specificity.

### Longitudinal Trajectories – Benign vs Malignant

We next examined how CYFRA 21-1 changed over time in each group. Figure 1 illustrates representative CYFRA trajectories (**Figure 1A**) for individual patients in the benign and malignant. Overall, patients with benign nodules showed relatively flat or gently increasing CYFRA 21-1 levels during follow-up, whereas the trends in cancer patients varied depending on treatment status. In the benign group, there was a slight upward drift in CYFRA values over time (many benign patients had very low values that rose by a small amount, though generally remaining in the normal range) (Figure 1B). The mixed-effects model quantified this: the *time* effect for the benign group was a positive but small slope (marginal mean of linear trend) (β = +0.00201on log(CYFRA) scale per day (95% CI: 0.00341 to 0.00369, p = 0.0185). In contrast, untreated cancer patients (those with cancer who had not yet been treated at the time of measurement) did not exhibit a significant increase over time (β = −0.00137, 95% CI: −0.00724 to 0.00450, p = 0.6468), implying a very slight decline, but with a wide confidence interval. Similarly, for the *treated-cancer* (post-surgery) group, the slope was β= –0.00263 (95% CI: −0.00797 to 0.00270, p=0.3326), a slight downward trend, but not significant (**Figure 1C**). In simpler terms, benign nodules showed a minor increase in CYFRA over time, whereas cancer patients (especially after tumor resection) tended to have stable or decreasing CYFRA levels over the follow-up period – but the variability was large, and these trajectory differences did not reach statistical significance in this sample.

**Figure 1.**
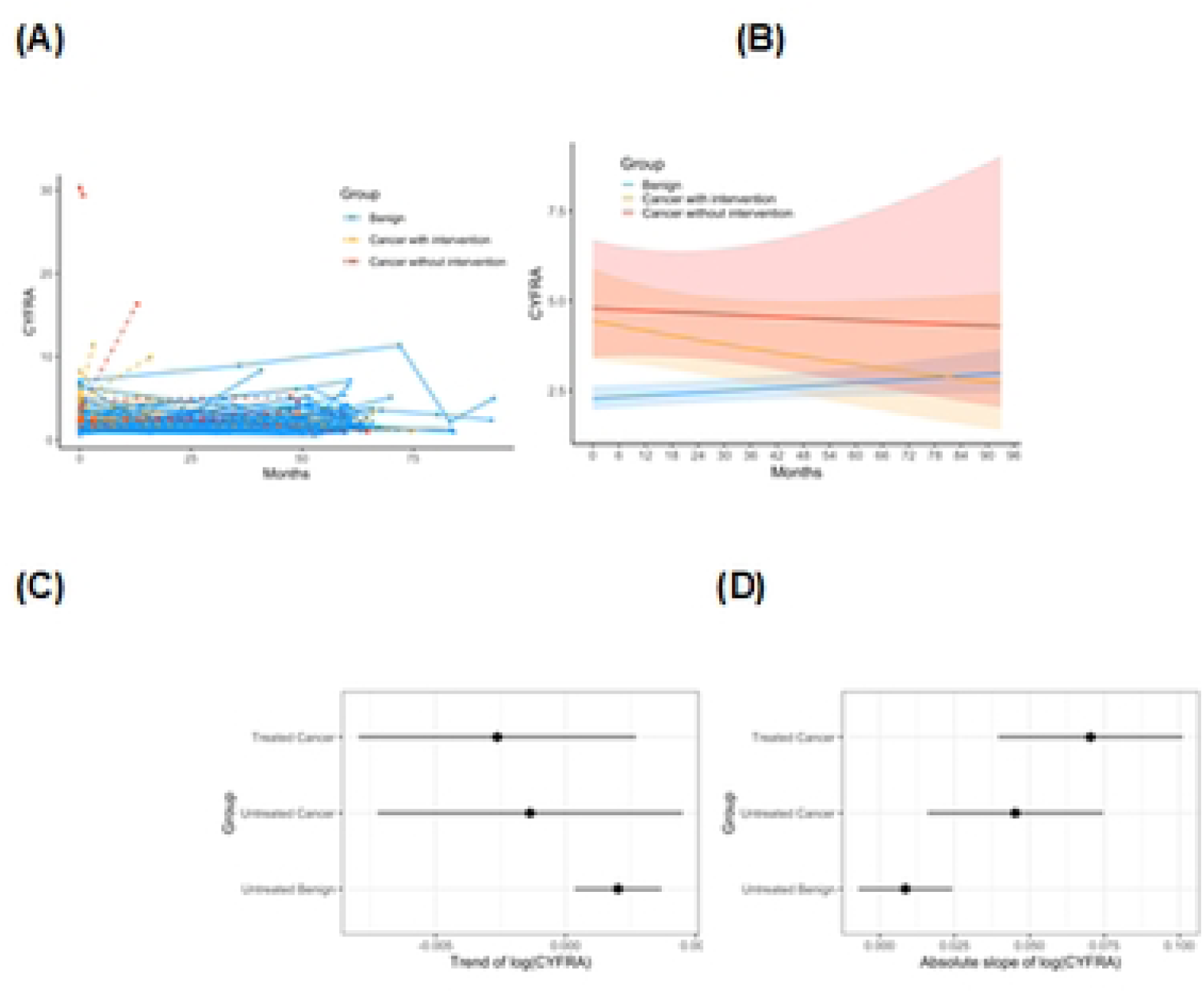
Longitudinal trajectories of CYFRA 21-1 in each patient (A), marginal means of overall trends by group in the mixed-effects model (B), marginal means of linear trend of log(CYFRA) by group based on mixed-effect model (C), marginal means of absolute slope of log(CYFRA) based on one-way ANOVA (D).

To assess differences in CYFRA 21-1 trajectories across diagnostic groups, the absolute slope of log-transformed CYFRA values was compared across groups. The ANOVA test showed the rate of changes in CYFRA over time differed significantly in at least one clinical group compared to the others (p = 0.0061). Estimated marginal means for the absolute slope of log(CYFRA) were 0.0086 (SE = 0.0080; 95% CI: −0.0071 to 0.0243) for benign,0.0453 (SE = 0.0137; 95% CI: 0.0159 to 0.0746) in pre/untreated cancer, and 0.0703 (SE = 0.0144; 95% CI: 0.0395 to 0.1012) for post treatment cancers (**Figure 1D**). Compared to the untreated benign group, both cancer groups demonstrated significantly higher absolute of CYFRA slopes. Specifically, untreated cancer patients exhibited a modest and increase in absolute slope (estimate = +0.0367, SE = 0.0158, p = 0.086) and treated cancer patients showed a larger increase (estimate = +0.0617, SE = 0.0164, p = 0.0028). However, the difference between treated and untreated cancer patients was not statistically significant (estimate = +0.0251, p = 0.368). These results suggest that CYFRA 21-1 levels change more rapidly over time in patients with malignancy, regardless of treatment status, compared to patients with benign nodules. Figure 1D displays the absolute slope of log(CYFRA) for each group with 95% confidence intervals.

### Impact of Surgical Treatment on CYFRA

Another way we evaluated this data was to consider patients before and after treatment separately, as some patients had several blood draws before treatment, after treatment, or both. For benign patients, we considered “Treatment” as the last date upon which we had a blood draw (**Figure 2A left panel**). For patients with cancer, we split blood draws into before and after (**Figure 2A right panel**). As expected, CYFRA 21-1 levels increase over time before treatment, seem to spike around the time of treatment, then decline after treatment.

**Figure 2.**
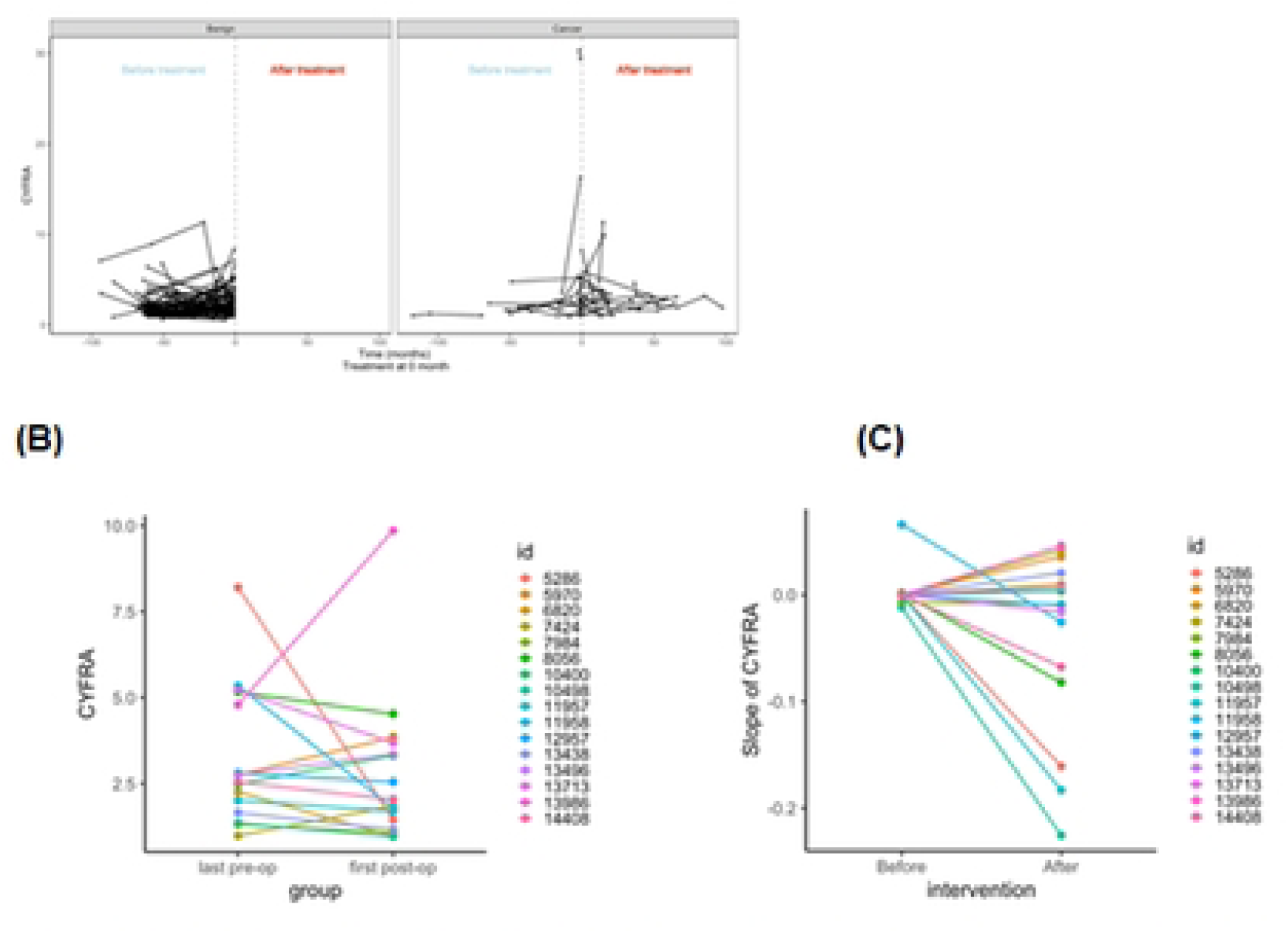
Trajectories tor Pre and Post treatment for benign(left) and cancer (right panel) (A), paired of before (the last pre-op measurement) and after (the first post-op measurement) treatment for 16 cancer patients (B), and paired slopes of before and after treatment for 16 cancer patients with longitudinal data before and after (C).

Focusing on the 16 lung cancer patients who had serial measurements both pre- and post-surgery, we directly assessed the effect of tumor resection on CYFRA 21-1 levels and trends. In the majority of these paired cases, CYFRA 21-1 decreased after surgery. The median CYFRA value before surgery (last pre-op measurement) was 2.64 ng/mL, which fell to a median of 1.94 ng/mL at the first post-op measurement in those patients (these values are illustrative; individual responses varied) (Figure 2B, p=0.433). Figure 2B showed that removal of the malignant nodule led to a prompt decline in the patient’s CYFRA 21-1, consistent with CYFRA originating from tumor activity. For example, one patient’s CYFRA dropped from 8.2 ng/mL before resection to 1.4ng/mL after surgery; another patient went from 1.35 ng/mL pre-op to 0.953 ng/mL post-op. This trend was not universal – a few patients had minimal change or even slight increases post-surgery (which could be due to post-operative inflammation or assay variation) – but the overall paired trend was a decline. These findings support that CYFRA 21-1 can serve as a dynamic marker of tumor burden: when the tumor is removed, the marker level falls correspondingly.

We then assessed the change in trends in patients before and after treatment in the 16 patients with matched samples. **Figure 2C** shows the paired change in slope for these patients. Prior to treatment, the slope of log(CYFRA) was minimal across patients (median = 0.00; mean ± SD = 0.00 ± 0.02), indicating relative biomarker stability during the untreated phase. After treatment, the slope became modestly negative (median = - 0.01; mean ± SD = −0.04 ± 0.08), suggesting a trend toward declining CYFRA levels post-intervention.

Despite this shift, the difference in slopes before and after treatment did not reach statistical significance (Wilcoxon signed-rank exact test, p = 0.211). These results suggest that although CYFRA levels tend to decline following surgical intervention, the rate of change across the post-treatment interval remains modest and variable among patients.

## Discussion

In this study, we investigated the longitudinal behavior of the serum biomarker CYFRA 21-1 in patients with pulmonary nodules, comparing those with benign versus malignant outcomes and evaluating changes in CYFRA after tumor removal. To our knowledge, this is one of the first analyses focusing on *within-patient CYFRA trajectories* over time. The results, albeit from a modest-sized cohort, suggest several important insights: (1) Lung cancer patients have higher CYFRA 21-1 levels than patients with benign nodules, not only at a single point (confirming known diagnostic patterns) but throughout the follow-up period. (2) The trajectory (slope) of CYFRA over time showed a tendency to differ by group (benign vs cancer), with benign nodules exhibiting a slight upward creep in CYFRA and cancers showing stable or downward trends (especially post-surgery), although this difference did not reach statistical significance in our sample. (3) Within individuals, effective treatment (surgical resection of a malignant nodule) was associated with a significant drop in CYFRA 21-1, reinforcing that CYFRA dynamically reflects tumor burden.

The finding that CYFRA 21-1 decreases after curative surgery is intuitively expected and aligns with prior observations that CYFRA can be used to monitor treatment response^12^. If a patient’s CYFRA remains high or rebounds after surgery, it could signal residual disease or early recurrence. This is analogous to the behavior of PSA in prostate cancer management: after prostatectomy, PSA should fall to nearly undetectable levels, and any subsequent rise indicates recurrence. CYFRA 21-1 could play a similar role in lung cancer surveillance. In our limited post-resection follow-up, none of the patients had known recurrence during the study period, and correspondingly their CYFRA stayed low after the initial post-surgery drop. Future studies with longer follow-up could assess whether rising CYFRA trajectories precede radiographic detection of recurrence.

The lack of a statistically significant difference in CYFRA *slopes* between benign and malignant groups in our analysis deserves discussion. We hypothesized that an actively growing cancer would show an increasing biomarker trajectory (analogous to a rising PSA in prostate cancer), whereas a benign nodule would not. However, several factors may have blunted our ability to see a clear divergence. Firstly, many cancer patients underwent surgery relatively early, truncating the “untreated tumor” observation period; their biomarker trajectory was reset by intervention (and we did observe the expected drop post-surgery). Secondly, some malignant nodules were indolent or treated with non-surgical therapies during follow-up (e.g., one patient received stereotactic radiation without resection) which could stabilize or reduce CYFRA levels, rather than showing unchecked increases. Thirdly, benign nodule patients, while lacking a tumor, are not a completely static group – slight CYFRA increases over time could result from aging or concomitant inflammatory lung conditions. Indeed, CYFRA 21-1, being a cytokeratin fragment, can be modestly elevated in certain benign lung diseases (e.g., chronic obstructive pulmonary disease or pulmonary fibrosis) and can fluctuate with acute exacerbations. Our benign group included a few patients with chronic lung conditions, which might explain the mild upward drift in CYFRA even without malignancy. Moreover, one benign patient had a transient extreme CYFRA elevation (11 ng/mL), likely due to an acute process; importantly, that level later normalized, and the nodule remained benign. Such cases illustrate how single high CYFRA readings can occasionally arise from benign causes – but if one tracks the trajectory, the benign cause tends to be self-limited (the biomarker spike resolved), whereas a malignant cause would be expected to produce a more sustained or rising pattern.

Our results underscore that *context and trajectory matter*: a CYFRA value moderately above the normal range in a nodule patient could be far more concerning if it is rising on serial tests than if it is stable or falling. This is precisely the lesson from decades of PSA-based prostate cancer screening – trends outperform one-off measurements in certain scenarios. In early lung cancer detection, recent evidence also supports leveraging serial measurements. For instance, a large study from the PLCO trial demonstrated that integrating biomarker trajectories (including CYFRA 21-1) improved lung cancer detection sensitivity while maintaining specificity^10^. Our work adds to this by focusing on the dichotomy of benign vs malignant nodules and by directly observing the effect of removing the tumor. We show proof-of-concept that CYFRA 21-1 trajectory analysis is feasible and can yield meaningful clinical signals (such as a post-surgery drop or an outlier rise in a “benign” case that warrants re-evaluation). In time, such longitudinal biomarker assessment could complement imaging in the management of indeterminate pulmonary nodules – for example, a steadily rising CYFRA might prompt earlier biopsy of a nodule that otherwise appears slow-growing on CT, whereas a flat CYFRA trend could support watchful waiting.

Several limitations of our study should be acknowledged. The sample size, especially of the malignant subgroup with long-term untreated follow-up, was small. Only a handful of cancer patients had prolonged observation without treatment, limiting our ability to detect a true difference in growth rates of CYFRA. Additionally, the follow-up duration was relatively short (most patients were followed for under one year), and thus we may have missed longer-term trajectory patterns (e.g., benign nodules might eventually plateau in CYFRA, or untreated cancers might eventually show exponential increases). The measurements frequency was not uniform – some patients had more frequent testing than others, which could influence the modeling (though our mixed-effects approach can handle irregular intervals, there could be bias if, say, sicker patients had more tests). We also did not account for potential confounders such as smoking status, histological subtype of lung cancer, or renal function (impaired renal clearance can elevate certain biomarkers); these factors could be explored in a larger cohort. Furthermore, while we centered our analysis on CYFRA 21-1, multiple biomarkers are often elevated in lung cancer. Some patients in our study also had elevated CEA, CA-125, etc., and a multi-marker trajectory might improve diagnostic accuracy over any single marker. Future studies could use panel approaches and advanced modeling (e.g., machine learning on serial data) to enhance discrimination of benign vs malignant nodules.

Despite these limitations, our findings have important clinical implications. They reinforce that a single high CYFRA 21-1 level in a nodule patient should be interpreted with caution, as it could either indicate cancer or be a false-positive from a benign cause; serial measurements can help tell these scenarios apart. If the CYFRA remains persistently elevated or trends upward, the likelihood of malignancy increases, whereas a downward or stable trend might favor a benign etiology or successful treatment. Additionally, monitoring CYFRA post-treatment can provide an early window into recurrence: even before a tumor is large enough to be seen on a scan, an upward inflection in the CYFRA trajectory might alert clinicians to microscopic residual disease. In this sense, CYFRA trajectory could become an adjunct “liquid follow-up” to radiographic follow-up.

### Conclusions

In summary, this longitudinal pilot study highlights that while CYFRA 21-1 has long been recognized as a lung cancer biomarker, examining its trajectory unveils new dimensions of its clinical utility. We observed that malignant pulmonary nodules are associated with higher and dynamically changing CYFRA levels, whereas benign nodules generally exhibit low and more stable CYFRA profiles. Though differences in rate-of-change between benign and malignant groups did not reach significance here, the trends suggest that with a larger sample or longer follow-up, such differences might emerge. The analogy to PSA dynamics in prostate cancer is apt – just as PSA velocity aids in prostate cancer decision-making, CYFRA 21-1 velocity or doubling time could enhance lung nodule risk stratification. We encourage further research with expanded cohorts to validate these findings. If confirmed, serial CYFRA 21-1 monitoring may become a valuable tool in the early detection of lung cancer and in the post-treatment surveillance for recurrence, ultimately improving clinical management of patients with pulmonary nodules.

## Supporting information

This work is supported by NIH National Cancer Institute [Grants 1R01CA253923 to F. M. and U01CA152662 to E. L. G.]

## Data Availability

The data underlying the results presented in this study are available from the Vanderbilt University Medical Center Institutional Review Board for researchers who meet the criteria for access to confidential data. Data cannot be shared publicly due to patient confidentiality and IRB restrictions (IRB# 030763). Request may be directed to the corresponding author or to the IRB office

## Acknowledgements

We would like to thank the clinical staff at Vanderbilt University Medical Center for their support in collecting and managing patient data. We also acknowledge the contributions of our biostatistical colleagues for their assistance with data analysis. Some sections of this manuscript were edited for grammar and style using AI-based language assistance tools.

## Notes

### Competing Interest Statement

The authors have declared no competing interest.

### Funding Statement

Yes

### Author Declarations

The study was conducted according to the Declaration of Helsinki in its current version and was approved by the Institution Board Review (IRB) of Vanderbilt University Medical Center (IRB#030763).

